# Investigating the role of whole genome sequencing in syphilis epidemiology: an English case study

**DOI:** 10.1101/2022.12.02.22283031

**Authors:** Mathew A. Beale, Louise Thorn, Michelle J. Cole, Rachel Pitt, Hannah Charles, Michael Ewens, Patrick French, Malcolm Guiver, Emma E. Page, Erasmus Smit, Jaime H. Vera, Katy Sinka, Gwenda Hughes, Michael Marks, Helen Fifer, Nicholas R. Thomson

## Abstract

**Background:** Syphilis is a sexually transmitted bacterial infection caused by *Treponema pallidum* subspecies *pallidum*, with approximately 6.3 million annual cases globally. Over the last decade, syphilis rates have risen dramatically in many high-income countries, including in England, which has seen a greater than 150% increase. Although this increase is known to be associated with high risk sexual activity in gay, bisexual and other men who have sex with men (GBMSM), cases are rising in heterosexual men and women, and congenital syphilis cases are now seen annually. The transmission dynamics within and between sexual networks of GBMSM and heterosexuals are not well understood.

**Methods:** To determine if whole genome sequencing could be used to identify discrete patterns of transmission, we linked national patient demographic, geospatial and behavioural metadata to whole *T. pallidum* genome sequences previously generated from 237 patient samples collected from across England between 2012 and 2018.

**Findings:** Phylogenomic analysis and clustering revealed two of the eight *T. pallidum* sublineages detected in England dominated. These dominant sublineages exhibited different spatiotemporal trends linked to demography or behaviour, suggesting they represent different sexual networks: sublineage 1 was found throughout England and across all patient groups, whereas sublineage 14 occurred predominantly in older GBMSM and was absent from samples sequenced from the North of England. By focussing on different regions of England we were able to distinguish a local heterosexual transmission cluster from a background of transmission amongst GBMSM.

**Interpretation:** These findings demonstrate that despite extremely close genetic relationships between *T. pallidum* genomes globally, genomics can still be used to identify putative transmission clusters for epidemiological follow-up, and therefore has a role to play in informing public health interventions.

**Funding:** Wellcome funding to the Sanger Institute (#206194 and 108413/A/15/D), UKRI and NIHR (COV0335; MR/V027956/1, NIHR200125), the EDCTP (RIA2018D-249), and UKHSA.

**Research in Context:** *Evidence before this study:* Detailed phylogenomic analyses investigating the epidemiology and transmission dynamics of *Treponema pallidum* are challenging due to low bacterial loads in clinical specimens, and difficulty in culturing the bacteria. We searched PubMed until August 9^th^ 2022 using the search terms “Syphilis” or “*Treponema pallidum*” and “genomic” or “genome(s)” or “sequencing”, finding 23 studies describing whole genome sequencing of *T. pallidum* subspecies *pallidum*, of which two used whole genome phylogenies to investigate sexual network epidemiology, with one large study of sexual networks conducted primarily in Victoria, Australia which characterised two major circulating sublineages in that setting, as well as putative sexual transmission networks with distinct sexual behavioural characteristics and potential bridging between networks.

*Added value of this study:* In this study, we linked national surveillance data to *T. pallidum* genomes, and characterised the transmission dynamics of syphilis using samples from across a whole country, in a European setting (England). Integration of national-level sociodemographic, spatiotemporal and genomic data allowed the delineation of putative sexual networks at both the national and region levels, and revealed patterns not previously detected using epidemiological or genomic data alone.

*Implications of all the available evidence:* Our findings are consistent with findings in Australia that demonstrate genomics can identify putative sociodemographic transmission clusters. However, in that study genomic clusters included samples separated by multiple single nucleotide polymorphisms, which could represent several years of evolution. Our study explored the value of linking identical genomes, and highlights that despite technical constraints, whole genome sequencing can be used to enable outbreak exclusion and identify putative local transmission clusters for epidemiological follow-up.

## Introduction

Syphilis is a sexually transmitted infection (STI) caused by the bacterium *Treponema pallidum* subspecies *pallidum* (TPA). Syphilis rates have been rising in many high income countries since the beginning of the 21^st^ Century^1–6^. In England (United Kingdom), new diagnoses of early syphilis (primary, secondary and early latent) rose from 3,011 (5.6/100,000 population) in 2012, to 8,011 (14.2/100,000 population) in 2019^4^. This increase has primarily been associated with gay, bisexual and other men who have sex with men (GBMSM) engaging in higher risk sexual behaviours^3,7,8^. However, cases amongst heterosexuals have also risen, raising concerns about infection during pregnancy and risks of vertical transmission leading to congenital syphilis^9,10^. Between 2016 and 2019, annual syphilis diagnoses in heterosexual men (MSW) and women (WSM) increased by 53% (660-1012) and 108% (294-614), respectively. There were 24 cases of congenital syphilis identified in England between 2015 and 2020, 15 of whom were born to mothers who tested negative at first trimester antenatal screening^11^, indicating they had acquired syphilis later in pregnancy. Some cases occurred in regions across England which had experienced recent increases in syphilis among women and GBMSM, suggesting that overlapping sexual networks may have facilitated wider dissemination^12^.

Although epidemiological surveillance provides insights into the current rise in syphilis rates, there are situations where this is insufficient. For example, a group of spatiotemporally clustered cases could represent a single outbreak and chain of transmission, but could also be the result of separate or unrelated transmission networks. Molecular typing methods^13–16^ provide one possible way to supplement epidemiological observations by identifying genetically related TPA strains. However, these methods may not accurately reflect recent evolutionary relationships between strains^17–19^, but instead cluster groups of bacteria which shared a common ancestor several decades ago, meaning it would be impossible to accurately delineate strain clusters relevant to epidemiologically useful timelines (usually months to years).

Whole genome sequence analysis (WGSA) has shown there are two co-circulating TPA lineages globally (Nichols or SS14) ^20–22^, which can be further divided into 17 sublineages plus singletons^20^. Further, these data have shown TPA genomes accumulate single nucleotide polymorphisms (SNPs) very slowly, with a median molecular clock rate equivalent to one substitution/genome every 6.9 years, meaning that isolate genomes from strains circulating in the UK can be identical (zero pairwise-single nucleotide polymorphisms (SNPs)) to those from Canada, Australia and other countries, and identical genomes were collected an average of 2.5 years apart (range 0-15 years). This genetic homogeneity has been suggested to indicate a recent global dissemination of TPA within the last 30 years, driven by a small number of multi-country sublineages^20^ and that genomic approaches may also be of limited value to investigate or resolve syphilis epidemiological links between patients.

To date, only a few studies have combined WGSA with patient demographic and sexual behaviour metadata to explore epidemiological trends of TPA, with studies from Japan^23^ and Australia^24^ finding discrete genetic clusters associated with GBMSM and heterosexuals. Here, we explored the value of WGSA for supplementing existing epidemiological data for understanding transmission at national and regional levels. We combined detailed patient demographic and epidemiological data with WGSA of TPA samples from England to gain insights into the different spatiotemporal and genomic transmission patterns of syphilis affecting GBMSM and heterosexuals.

## Methods

### Samples and patients

A detailed description of sample collection and patient metadata linkage is provided in the Supplementary Methods and is summarised in Supplementary Figure 1. Briefly, TPA positive genomic DNA samples were retrieved from historical archives (2012-2017) held at the UK Health Security Agency (UKHSA, previously Public Health England) National Reference laboratory for bacterial STIs (Colindale, North London), as well as being prospectively collected (2016-2018) from five laboratories (Birmingham, Brighton, Leeds, London (University College London Hospitals/Mortimer Market Clinic), Manchester) with high syphilis case-loads who perform in-house molecular TPA diagnostic testing (and thus do not usually refer to the UKHSA reference laboratory).

For samples from UKHSA, patient metadata were obtained by linkage to the National STI surveillance system (GUMCAD). For samples prospectively collected from the five non-referring laboratories, patient metadata available from local laboratory information systems was linked to GUMCAD data and integrated into the larger dataset after deduplication (see Supplementary methods and Supplementary Figure 1). For comparison between the sequencing dataset and national surveillance rates, we also retrieved summary statistics from GUMCAD data for all syphilis patients 16 years and older in England from 2012-2018 (n=50,845).

### Sequencing and phylogenomic analysis

Whole genome sequencing of all clinical *T. pallidum* samples used in this study has been previously described^20^, and was performed directly on the residual genomic DNA extracts from residual diagnostic samples using the pooled sequence capture method^21^ on Illumina HiSeq 4000. Genomic data were analysed as previously (see Supplementary Methods).

### Ethics and data governance

Ethical approval for all clinical samples was granted by the National Health Service (UK) Health Research Authority and Health and Care Research Wales (UK; 19/HRA/0112) and the London School of Hygiene and Tropical Medicine Observational Research Ethics Committee (REF#16014). Diagnostic samples were identified at UKHSA using internal laboratory information systems. UKHSA has permission to process confidential patient data under Regulation 3 (Control of Patient Information) of the UK Health Service Regulations 2002. Information governance advice and ethics approval for this study were granted by the UKHSA Research Ethics and Governance Group. Full details of approvals and pseudonymisation of samples and patient metadata are described in the Supplementary Methods.

## Results

### Demographics of syphilis in England

Of 497 English samples submitted for sequencing, we recovered 240 high quality genomes (198 from the UKHSA reference laboratory, 42 from other laboratories; Supplementary Figure 1). Three duplicate samples (same patient and collection date) were included in the main phylogenies, but removed from further analyses of English populations, leaving 237 genomes. Two hundred and seventeen samples were grouped into nine official UKHSA Regions; for 18 samples the region was unknown; two samples were referred to English laboratories from elsewhere in the UK (Supplementary Figure 2). Our samples were dominated by those from London (n=118), with the South East (n=29), North East (n=24) and South West (n=15), the next largest groups. Analysis of collection dates showed that although we had sequences from most regions throughout the study period, the North West and Yorkshire and Humber were represented only at the beginning (2012, 2013) and end (2018) of the timeline.

Across the national genome collection, 76.0% (n=180) came from GBMSM, compared to 10.6% (n=25) MSW, 6.3% (n=15) men with unrecorded sexual orientation (MUnknown), 3.8% (n=9) WSM, and 3.4% (n=8) Unknown gender and sexual orientation (Supplementary Figure 2). Notably, the most highly represented region (London) had a higher proportion of GBMSM (93.2%, n=110) compared to the next most highly represented regions (South East, 72.4%, n=21; North East, 62.5%, n=15; South West, 80.0%, n=12). Due to a low number of heterosexual individuals in the UKHSA dataset, HIV status was restricted to GBMSM to prevent deductive disclosure (for the prospectively collected samples, there were no MSW/WSM living with HIV). Within the genome dataset, 27.4% of cases were living with HIV, and these were distributed across seven out of nine regions, with London, containing mostly GBMSM, having the highest proportion (n=48, 40.7%).

To establish how representative our genome collection was of the UK syphilis cases in England, we compared the distributions of socio-demographic characteristics of cases in the WGS dataset with those from the national STI surveillance system (GUMCAD) (Supplementary Table 1). Overall, our WGS sample dataset (n=237) represented 0.5% of all syphilis diagnoses among patients 16 years and older in England during the period 2012-2018 (n=50,845). Compared to diagnoses reported in GUMCAD, the samples used in the WGS project were broadly representative by age group, region of residence (including London vs non-London), and HIV status (GBMSM only). However, a higher proportion of cases in the WGS dataset were GBMSM (75.9% vs 65.3% respectively), with fewer women (3.8% vs. 12.7% respectively) and heterosexual men (10.5% v.s. 17.7%). The WGS dataset also had a much higher proportion of primary syphilis cases compared to GUMCAD (81.4% compared to 27.9% respectively), largely reflecting the clinical presentation of primary syphilis with ulcers which permit swabbing.

We inferred the presence of macrolide resistance conferring SNPs in the ribosomal 23S as previously described^21^, and found that 88.2% of UK genome samples carried the A2058G allele, 2.5% had an uncertain or mixed variant call at position 2058, and 2.1% carried the A2059G allele, meaning that only 7.2% of English TPA genomes carried a wild type ribosomal 23S gene and were therefore predicted to be sensitive to macrolide antimicrobials.

### Genomic clustering of samples

A whole genome phylogeny was inferred from the 237 English genomes sequenced here, along with 286 global contextual genomes. We clustered all isolates by lineage, sublineage, or into single-linkage SNP clusters. The English genomes were broadly distributed throughout the known TPA phylogeny (Supplementary Figure 3). Referencing previous work by us and others^20–22^, 77.2% (n=183) of these genomes belonged to SS14-lineage and 22.8% (n=54) to the Nichols-lineage. Of the seventeen defined global sublineages^20^, eight were present in the UK, along with one singleton (Figure 1A, 1B, 1C, Supplementary Figure 4A). The English samples were dominated by the global sublineages 1 (n=175) and 14 (n=44), but two other globally distributed sublineages (2, n=5; 8, n=5) were also detected in the UK (Figure 1A, 1C) as well as two English samples for each of sublineages 3, 6 and 15, and one for sublineage 16.

**Figure 1.**
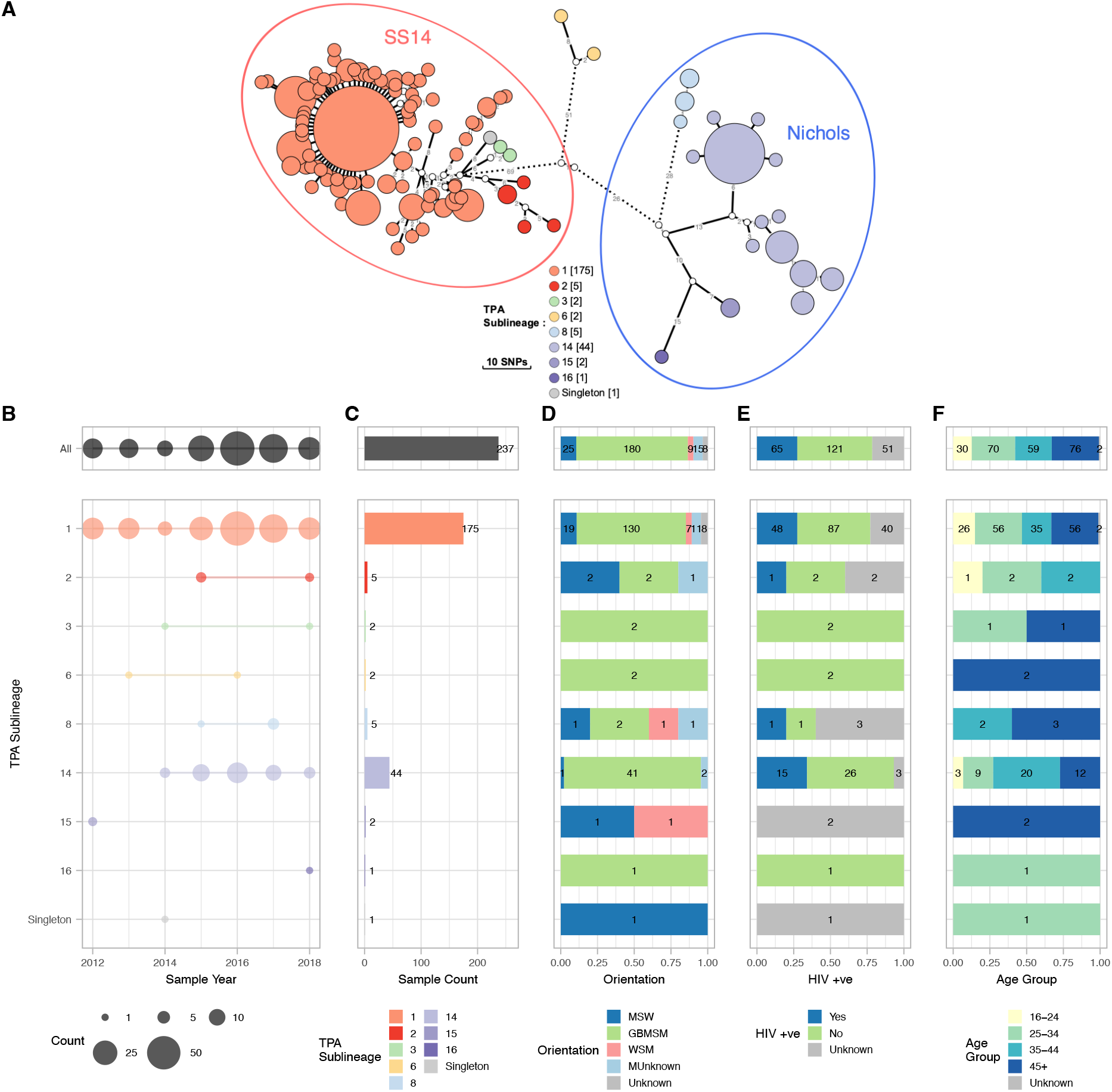
Population structure of English *T. pallidum* genomes according to phylogenetic sublineages, and associated patient characteristics. A – Minimum spanning tree visualisation of genetic relationships between samples from England. Node size corresponds to the number of identical genome samples in a cluster, and edge length (with number) to the number of substitutions between identical genome sample clusters (where edges were longer than 12 substitutions, these have been shortened - indicated by dashed lines). Nodes are coloured according to the TPA sublineages defined previously^20^, and in the legend, numbers in parentheses indicate total sample count for each sublineage. Primary Lineage (SS14/Nichols) is indicated by encompassing ellipses; sublineage 6 diverges from other TPA close to the root, and has previously been classified as Nichols^20^. B – Samples per collection year per sublineage. C - Total sample counts per sublineage. Bar plots show proportion of each group by D - Gender Orientation, E - HIV Status, F - Age Group (numbers indicate exact sample counts).

Linking previously sequenced whole TPA genomes from English patients to STI surveillance and laboratory records, we observed that patients infected with the most common sublineage (sublineage 1; 175/237, 73.8%) were largely representative of syphilis patients overall, with 74.3% classified as GBMSM (130/175) (Figure 1D, Supplementary Figure 4B) and 52.0% (91/175) aged 35 or older. In contrast, 93.2% (41/44) of patients infected with sublineage 14 were GBMSM (1/44 MSW, 2/44 MUnknown), significantly more than would be expected by chance (Fisher’s Exact test, p=0.0087); 73% (32/44) were age 35 or older, and 34.1% were living with HIV. We also found that most patients living with HIV were infected with sublineages 1 or 14, consistent with these lineages being linked to GBMSM networks (Figure 1E, Supplementary Figure 5). While seven women (4.0%) were infected with TPA sublineage 1, there were no women infected with sublineage 14 (Figure 1D, Supplementary Figure 4B). We found some rarer sublineage groups contained a higher proportion of heterosexual men and women, with sublineage 8 containing two heterosexuals (1/5 WSM, 1/5 MSW), whilst sublineage 15 comprised two heterosexuals (1/2 WSM, 1/2 MSW) residing in the East of England. Analysis of the genetic relationships indicated that, at least in these two examples, the heterosexual samples were genetically identical to one another but distinct from other samples, falling on terminal nodes of our minimum spanning network (Supplementary Figure 4B).

To explore this further, we delineated genomes from English patients into 27 distinct clusters of two or more genomes with zero pairwise-SNPs between them (i.e. identical at the core genome level; Supplementary Figure 6). Given the genetic homogeneity of TPA, these clusters do not necessarily indicate direct patient-to-patient transmission, but instead provided a means of clustering samples sharing a recent common ancestor. Of the 146/237 genomes included in these zero-distance genome clusters, 20 were from patients identifying as heterosexual (MSW, n=15; WSM, n=5), and 11/20 (55%) of these heterosexuals were part of a cluster containing only other heterosexual individuals. In particular, of the four genome clusters containing WSM, three of those only contained other heterosexuals and each of these clusters was detected in only a single region. However, the fourth cluster containing a WSM was the largest cluster of identical genomes, comprising 42 samples from sublineage 1, of which 35 patients (83%) were GBMSM, compared to 3 MSW, 1 WSM, and 3 with unknown sexual orientation. Therefore, this primarily GBMSM cluster also includes four patients who identify as heterosexual, indicating there may be some bridging between the populations.

In our previous global analysis, we identified two samples from England which formed sublineage 6, an unusual phylogenetic outgroup close to the most recent common ancestor (MRCA) of all TPA. To understand the origins of this sublineage, we interrogated the newly available patient metadata for the samples, and found both were from HIV negative or undiagnosed GBMSM aged over 45. The first sample (PHE130048A) was collected in 2013 in London from a man born outside of the UK, while the second (PHE160283A) was collected 3 years later in 2016 in the South East of England from a UK-born man (no further information was available regarding travel, immigration history, or epidemiological linkage between the patients). We examined the pairwise-SNP distances between these two samples, and found them to be separated by 14 pairwise-SNPs, with PHE160283A separated by 8 SNPs from the MRCA of the two samples. Given both the genetic and temporal distances between the two samples, they were highly unlikely to be directly related to one another in a chain of transmission, and therefore could either represent sampling of a rare lineage circulating in the UK, or two separate introductions from another country where this lineage is more common; this has not been described elsewhere, but is plausible given the low depth of sampling for most countries.

To understand the global context of TPA from England, we generated a Bayesian time-scaled phylogeny using the global dataset (Supplementary Figure 7; comprising contextual genomes from 21 other countries), and examined subtrees for the four most common English sublineages (1, 2, 8, 14; Supplementary Figure 8). Sublineage 1, previously found to be globally disseminated^20^, contained samples from around England and the world; the English samples were polyphyletic and distributed throughout the Sublineage 1 phylogeny, with only limited clustering (Supplementary Figure 8A), with the exception of a clade predominantly comprising samples from North East England (see below). Consistent with previous observations, 89.7% of sublineage 1 strains carried the ribosomal 23S A2058G allele, and were predicted to be resistant to macrolides^20,21^ (Supplementary Figure 9). As previously described^20^, sublineage 2 comprises two clades, one of which is dominated by North American samples, with the other dominated by samples from China. English samples were found within both clades, with at least three distinct groupings of English strains (Supplementary Figure 8B), likely indicating multiple recent independent introductions from other countries. In contrast, we found five sublineage 8 samples from England forming a monophyletic subclade along with individual samples from Canada and Australia (Supplementary Figure 8C). Sublineage 14 represented a major English sublineage, with 44/55 sublineage 14 genomes in the global collection coming from England, all of which had the ribosomal 23S A2058G allele. We previously described the contemporaneous appearance of this sublineage in England and Canada in 2013/2014^20^, but our time-scaled phylogeny shows two clades within sublineage 14, both of which have median time to MRCAs (1999 and 2006) predating the first detection in our dataset, suggesting this sublineage had been circulating in England for some time (Supplementary Figure 8D).

### Investigation of regional differences in population structure

We examined the geographical distribution of types in England and found that both SS14- and Nichols-lineages were co-circulating in London and throughout South and Central England (Figure 2A, 2E). However, we found only SS14-lineage in the three most northerly regions (North East, North West, Yorkshire and Humber) (Figure 2A, 2E). Examination of TPA sublineage distributions indicated that sublineage 1 (SS14-lineage) was present in all regions (and represented the only sublineage present in the three northern regions), whilst sublineage 14 (Nichols-lineage) was co-circulating with sublineage 1 in London, the South and Central England but not in Northern England (Figure 2B, 2F). There were 35 samples from the three northerly regions, of which 49% (17/35) were collected after the first detected appearance of sublineage 14 in 2014, coinciding with an increase in national syphilis rates^25^. Since prevalence of sublineage 14 within the overall dataset was 18.6%, its absence from samples in the northern region could suggest different patterns of transmission.

**Figure 2.**
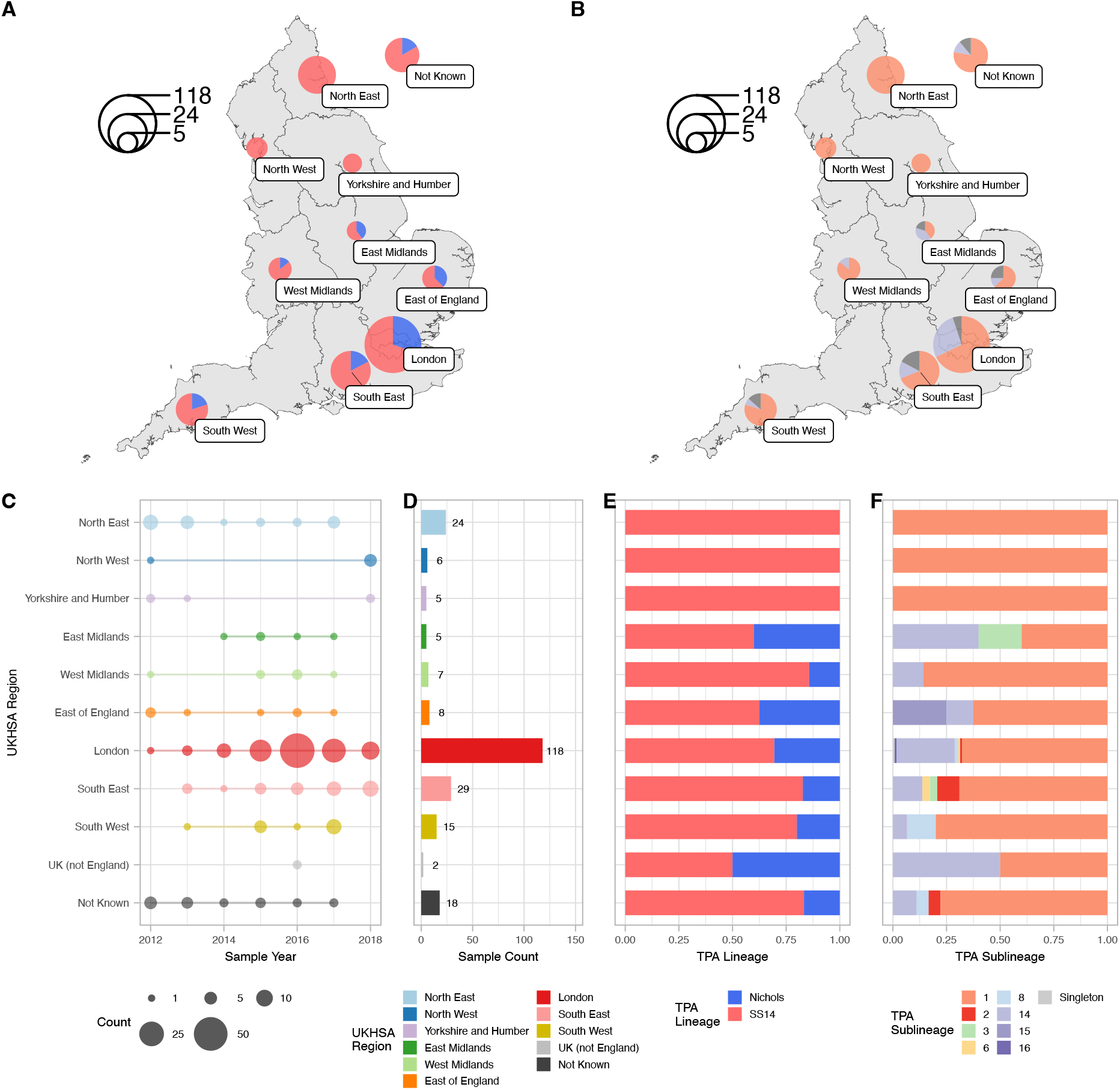
Geographical distribution of English genome samples and according to phylogenetic sublineages. A- Map showing proportion of samples from each UKHSA Region of England by TPA Lineage (Red=SS14, Blue=Nichols). B- Map showing proportion of samples in each UKHSA region of England for the two most common TPA Sublineages (Orange=Sublineage 1, Blue=Sublineage 14, Grey=Others). C- Distribution of sample collection years, D- total sample counts, E- proportion of samples from each region each by Lineage, F- proportion of samples from each region by Sublineage (numbers in bar plots indicate exact sample counts)

Apart from the three northern regions and the West Midlands (which contained only sublineages 1 and 14), all other regions contained at least three sublineages (range 3-7). London represented 49.7% (118/237) of samples in the study, and here we detected six sublineages (1, 2, 6, 7, 14, 16) and one sublineage previously labelled as a singleton. As elsewhere, London was dominated by sublineages 1 (n=80, 67.8%) and 14 (n=32, 27.1%). The rare sublineages 15 (n=2) and 16 (n=1) were each found in a single region (East of England and London respectively), but all other sublineages were found in multiple regions (Figure 2).

Our geospatial analysis showed that all samples from the three most northerly regions of England were from SS14 sublineage 1 (Figure 2), contrasting with greater diversity elsewhere in England. To explore this in more detail, we focussed on the 24 samples collected from the North East of England between 2012 and 2017. Although all samples belonged to sublineage 1, grouping North East samples using a two pairwise-SNP threshold identified three distinct North East clusters within sublineage 1 (Figure 3A), and correlation with the global phylogeny (Figure 3B, Supplementary Figure 8A) indicated different distributions (Figure 3B). Cluster 1 comprised samples from 13 cases collected between 2012 and 2017, the majority (12/13) were GBMSM. Within this cluster, samples could be further subdivided into three subgroups of eleven samples linked by identical core genomes (zero pairwise-SNPs; Figure 3A). Phylogenetic analysis of all English and global samples from sublineage 1, showed that although posterior support for internal nodes was low, the North East samples appeared to be polyphyletic (Figure 3B), and interspersed with samples (including additional identical genomes) from the rest of the England, and around the world. Therefore, it is unlikely that the North East samples from Cluster 1 represent a direct chain of transmission or local outbreak, but rather that we have sampled from a broader transmission network spanning national and global boundaries.

**Figure 3.**
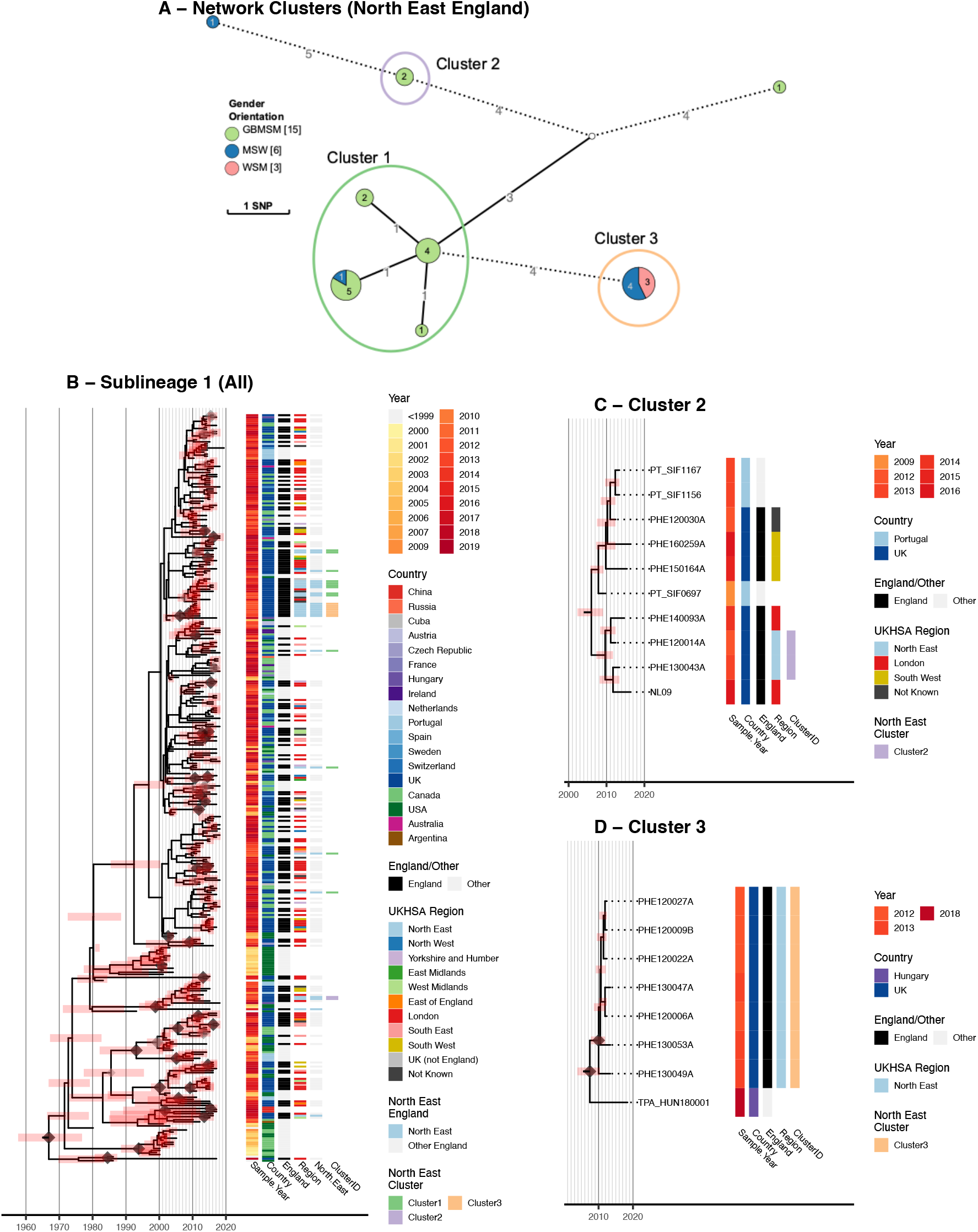
Spatiotemporal and genomic clustering analysis of samples from North East England. Sublineage 1 samples from North East England (22/24) formed three distinct clusters. A- Minimum spanning tree visualisation of genomic relationships between samples in North East England. Node size corresponds to the number of identical samples, and edge length (with number) to the number of substitutions between clusters (where edges were longer than 3 substitutions, these have been shortened, indicated by dashed lines). Nodes are coloured by proportion of patient gender orientation (MSW, Men who have sex with women; GBMSM, Gay, bisexual and other men who have sex with men; WSM, Women who have sex with men). Clusters were defined by connections to another sample within 2 pairwise-SNPs. Clusters 1 and 2 appear to be dominated by GBMSM populations, whilst Cluster 3 contains only patients identifying as heterosexual. B- Time-scaled sublineage 1 tree of global samples indicates that the GBMSM-associated Cluster 1 is a globally distributed cluster, and that the North East samples are polyphyletic. C- Time-scaled subtree of global samples sharing a common ancestor with Cluster 2 indicates a close relationship with two samples from London, and more distantly with those from South West England and Portugal. D- Time-scaled subtree of global samples sharing a common ancestor with the heterosexual-associated Cluster 3 suggests the North East England samples are closely related to each other, but not to any others, with the closest related strain found in Hungary. This could imply a historic importation from another country, followed by local circulation.

Cluster 2 comprised two samples collected in 2012 and 2013 from GBMSM with identical core genomes, and similarly to Cluster 1, formed a clade with eight other samples, of which five were from elsewhere in England (4/5 GBMSM, 1/5 Unknown) and three were from Portugal^26^ (Figure 3C). In contrast, Cluster 3 comprised seven samples which all came from the North East in 2012 and 2013, and had identical core genomes (zero pairwise-SNPs). These samples formed a monophyletic clade, with the nearest related other sample in the global dataset separated by two SNPs and isolated from a bisexual man in Hungary in 2018^20^ (Figure 3D). All seven Cluster 3 patients self-identified as heterosexual (MSW or WSM). Given the close spatiotemporal and genomic relationships between Cluster 3 samples, and contextualised by a background of greater diversity over the 2012-2017 timespan of all other samples collected from North East England, Cluster 3 likely represents a localised outbreak in a heterosexual network. Indeed this observation, made solely on the basis of the available genomic and sociodemographic data, is consistent with reports of a syphilis outbreak occurring amongst heterosexuals in the North East^27^. It is likely that some of our samples are derived from this event.

## Discussion

In this study, we linked patient demographic, geospatial and behavioural metadata, to previously generated TPA genomes from 237 patients diagnosed in England between 2012 and 2018. Our analysis shows a variety of English sublineages, dominated by global sublineages 1 and 14^20^, both of which are predicted to be resistant to macrolides and consistent with the high percentage of macrolide resistant samples in the UK. The English sublineages 1 and 14 displayed different patient sociodemographic and spatiotemporal profiles, with sublineage 1 patients showing a greater diversity of gender, sexual orientation, HIV status and age, while sublineage 14 was primarily found in older GBMSM. Moreover, whilst sublineage 1 was found in all regions of England, cases attributed to sublineage 14 were mainly taken in London, and not found in the northern regions of England. These contrasting characteristics suggest that the two sublineages describe distinct sexual transmission networks, consistent with a recent WGSA study from Australia^24^, which identified broadly similar TPA population structures co-circulating in Melbourne and the Northern Territory. Notably, both common sublineages (1 and 14) contained both HIV positive and negative cases, and there was no phylogenetic delineation by HIV status, suggesting either that HIV status may not be strongly associated with transmission patterns, or that such patterns are beyond the ability of WGS-based analyses to detect.

We were also able to examine whether the data could be used to define GBMSM and heterosexual transmission networks based on the proportion of individuals identified as GBMSM or heterosexual for each genomic cluster^24^. We observed three instances where genomes from heterosexual individuals clustered with identical genomes only from other heterosexuals from the same region, consistent with this representing discrete heterosexual transmission networks or clusters. Contrastingly, we found that many genetic clusters classified as GBMSM-associated under a proportional definition across the whole dataset exhibited spatiotemporal diversity. We also found mixed clusters, in particular a large cluster of 42 samples with identical core genomes, the majority from GBMSM individuals, four from heterosexuals (one woman, three men) and three with unreported sexual orientation and/or gender. Samples in this cluster had diverse regional geography and spanned across the 7 years of this study, and this implies widespread dissemination through the population more rapidly than the bacteria acquires variation, and potentially represents multiple local transmission networks all sharing a recent common ancestor. The presence of heterosexuals within these networks indicates possible bridging between transmission network groups^28^.

As in most countries, sublineage 1 dominated among samples from England^20,24^. Whilst the majority (74.3%) of English sublineage 1 patients were GBMSM, with TPA genomes occupying positions across the sublineage 1 phylogeny and interspersed with samples from around the world, in the North East of England we identified a genetically distinct cluster of identical core genomes found exclusively in heterosexuals, consistent with reports of a syphilis outbreak amongst heterosexuals at that time^27^. Given the previous uncertainty as to whether genomics can play a substantial role in understanding the epidemiology of syphilis due to TPA’s genetic homogeneity and low molecular clock rate^20,24,29^, our work suggests WGSA combined with detailed epidemiological data can resolve some local transmission chains for TPA. This could offer opportunities to intervene or educate regarding particular sexual networks, as well as to determine outbreak membership.

In other STIs such as *Neisseria gonorrhoeae*, SNP cut-offs of either 5 or 10 SNPs have been used to infer transmission chains^28,30^. *N. gonorrhoeae* accumulates SNPs at a rate of 8 substitutions/genome/year^31^, nearly 60 times faster than TPA. Therefore, even TPA isolates with identical genomes do not necessarily indicate recent direct patient-to-patient transmission; conversely samples separated by even a very small number of SNPs are unlikely to share a recent common ancestor. Furthermore, because the potential transmission window of TPA may be as high as two years^32,33^, direct transmission cannot be excluded temporally for identical genomes collected within that period. This may ultimately limit our overall ability to deconvolute national/regional scale patterns of transmission.

This collection of genomes represented 0.5% of the recorded number of syphilis cases in England during the study period and all samples referred to the National Reference laboratory with sufficient Treponemal DNA for sequencing. Whilst the available referral population may not be fully representative of syphilis in England due to regional variation in molecular testing and referral practices, all samples were collected and sequenced in the absence of any genetic relatedness information, so our genomic observations provide a snapshot of circulating English lineages. Future studies which focus on the systematic collection of samples from a higher proportion of cases, combined with improved sequence quality will enable further insights into TPA transmission dynamics, and enable the fuller utility of sequence data to inform public health interventions.

## Data Availability

Sequencing reads for all genomes used in this study have been previously published and described, and are available at the European Nucleotide Archive (https://www.ebi.ac.uk/ena) in BioProjects PRJEB28546, PRJEB33181 and PRJNA701499. All accessions, corresponding sample identifiers and related metadata are available in Supplementary Data 1. Patient metadata for the UK genomes is available in pseudonymised form in Supplementary Data 2. UK shape files for Public Health (UKHSA) region boundaries were downloaded from the UK Office for National Statistics, available at https://geoportal.statistics.gov.uk. All sample metadata and intermediate analysis files are available at https://doi.org/10.6084/m9.figshare.21543333.v1 and https://github.com/matbeale/Syphilis_Genomic_Epi_England_2022-23.

https://github.com/matbeale/Syphilis_Genomic_Epi_England_2022-23

## Code Availability

The R code for all phylogenetic and statistical analysis and plotting is available in an Rnotebook at https://doi.org/10.6084/m9.figshare.21543333.v1 and at https://github.com/matbeale/Syphilis_Genomic_Epi_England_2022-23, along with underlying source files.

## Author Contributions

Conceptualisation: MAB, MJC, GH, MM, HF, NRT; Methodology: MAB, LT; Formal Analysis: MAB, LT; Investigation: MAB, LT, MJC, RP, HC; Resources: MJC, RP, ME, PF, MG, EEP, ES, JHV, KS, GH, MM, HF; Data Curation: MAB, LT, HC, KS; Writing – Original Draft: MAB; Writing – Review & Editing: MAB, LT, MJC, RP, HC, KS, GH, MM, HF, NRT; Visualisation: MAB; Supervision: MJC, KS, HF, NRT; Project Administration: MAB, MJC, MM; Funding Acquisition: KS, MM, HF, NRT.

## Acknowledgements

We thank the sequencing team at the Wellcome Sanger Institute, and C. Puethe and the Pathogen Informatics team for computational support, and additional technical staff involved in sample diagnostics, DNA extraction and sample retrieval in laboratories at Public Health England (now UKHSA) and NHS laboratories (Birmingham, Brighton, Manchester, Leeds, London (UCLH)), UK. MAB and NRT were supported by Wellcome funding to the Sanger Institute (#206194 and 108413/A/15/D). MM was funded by the UKRI and NIHR (COV0335; MR/V027956/1, NIHR200125) and the EDCTP (RIA2018D-249). Staff time for LT, MJC, RP, HC, KS, HF, patient metadata retrieval and analyses were funded internally by UKHSA.

This research was funded in whole, or in part, by the Wellcome Trust (#206194 and 108413/A/15/D). For the purpose of open access, the authors have applied a CC-BY public copyright licence to any author-accepted manuscript version arising from this submission.

