## Supplementary Information for "Investigating the role of whole genome sequencing in syphilis epidemiology: an English case study"

**Beale et al, 2022**

**Supplementary Information**

### Supplementary Methods

#### Samples and patients

A diagrammatic representation of the sampling and metadata linkage framework is shown in Supplementary Figure 1. The UK Health Security Agency (UKHSA) National Reference laboratory for STIs (Colindale, North London) holds a collection of samples of syphilis-positive ulcer swabs from 2003. Records for 1,139 TPA PCR positive samples archived at UKHSA (2012-2017) were retrieved and qPCR results analysed. Previous work has shown that pathogen load is a key determinant for success in recovering whole TPA genomes[1,2]. Six hundred and ninety samples with a historic qPCR Cq <34 were recovered from freezer archives and shipped to the Wellcome Sanger Institute (WSI; Cambridgeshire), where all qPCRs were repeated using a Tp47 specific amplicon as previously described[1]. 434 samples with a repeat Cq < 33.6 were submitted for whole genome sequencing.

In addition, 173 samples were prospectively collected (2017-2019) from five laboratories (Birmingham, Brighton, Leeds, London (University College London Hospitals/Mortimer Market Clinic), Manchester) with high syphilis case-loads who perform in-house molecular TPA diagnostic testing (and thus do not usually refer to the UKHSA reference laboratory). Notably, for some cities (e.g. London) and all regions, we had samples referred from these specific laboratories as well as within the UKHSA collection, meaning regional data was captured by both datasets. All samples were screened by qPCR at WSI, and following evaluation of sequencing performance from the UKHSA samples, 63 samples with a more conservative qPCR threshold of Cq <32 were submitted for sequencing.

For 185 samples from UKHSA, laboratory records for were linked to the National STI clinical surveillance reporting database (GUMCAD) using internal patient identifier, and cross-checked for accuracy using clinic attendance date and sample receipt date. A further 10 samples from UKHSA did not match to a record in GUMCAD, however the sample included data on age and whether the patient lived in London or the rest of the UK. Duplicates and quality controls (n=7) were removed from the analysis. Where the patient identifier matched to multiple GUMCAD attendances, sample receipt date was used to select the correct attendance date. Attendances for a syphilis diagnosis that were within 60 days before or after the receipt date were selected, with an attendance date before the receipt date prioritised. If there was no attendance for diagnosis, attendance for a syphilis test that was within 60 days before or after the receipt date was chosen. Manual deduplication was required for patients that had no attendance or test date close to the sample receipt date. Syphilis stage information was only available for samples matched to a syphilis diagnosis (n=140). For comparison between the sequencing dataset and national surveillance rates, we also retrieved summary statistics from GUMCAD data for all syphilis patients 16 years and older in England from 2012-2018 (n=50,845). The final linked dataset was limited for patient confidentiality purposes, and included information on the year and month (where available – for a small number of samples where month was unknown, this was set to June) of sample collection, English region ("PHE centre"), patient gender and sexual orientation, patient age group, HIV status (for GBMSM patients only, to prevent deductive disclosure in heterosexuals), and whether the patient was born in the UK.

Patient metadata for samples prospectively collected from the five laboratories in Brighton, Birmingham, Leeds, London and Manchester were retrieved from local laboratory information systems. In most cases, we were able to access details on patient gender, declared sexual orientation, HIV status (for GBMSM patients and a subset of others), age, geographical region, and sample collection year. We also searched for samples linked to the same patient identifiers to enable deduplication across the full dataset. We integrated the patient metadata from these samples into the dataset retrieved by UKHSA from GUMCAD. A full description of sample and data flow is shown in the CONSORT diagram from Supplementary Figure 1.

#### Ethics and data governance

Ethical approval for all clinical samples was granted by the London School of Hygiene and Tropical Medicine Observational Research Ethics Committee (REF#16014) and the National Health Service (UK) Health Research Authority and Health and Care Research Wales (UK; 19/HRA/0112). Diagnostic samples meeting selection criteria were identified at UKHSA using internal laboratory information systems (LIMS). Samples had no patient identifiable information other than an internal sample number, and were transferred to the WSI for testing and whole genome sequencing, initially linked to sample collection year only. UKHSA has permission to process confidential patient data under Regulation 3 (Control of Patient Information) of the Health Service Regulations 2002. Information governance advice and ethics approval for this study were granted by the PHE (now UKHSA) Research Ethics and Governance Group. Limited patient metadata was retrieved from GUMCAD, by UKHSA, and used to generate summary statistics about populations. Linkage of patient metadata to samples was performed at UKHSA, and was fully pseudonymised before transfer to WSI, where it was held on an encrypted server with access restricted to study investigators. Heterosexual individuals who were also HIV positive were expected to be rare in the dataset, and to minimise the risk of deductive disclosure of patient identity, HIV status was not provided for these individuals. Collection and sharing of residual diagnostic samples and patient metadata prospectively collected by non-referring laboratories were included within the sample ethics described above. Patient metadata for these samples was retrieved from hospital LIMS by the local investigator, pseudonymised using a key held only by the referring laboratories, before transfer to WSI.

#### Sequence analysis

Whole genome sequencing of all clinical *T. pallidum* samples used in this study has been previously described[3], and was performed directly on the residual genomic DNA extracts from residual diagnostic samples using the pooled sequence capture method[1,4] on Illumina HiSeq 4000. Sequencing reads were filtered using the full bacterial and human Kraken 2[5] v2.0.8 database (March 2019) to extract *Treponema* genus-specific sequencing reads, followed by trimming with Trimmomatic[6] v0.39 and downsampling to a maximum of 2,500,000 using seqtk v1.0 (available at https://github.com/lh3/seqtk) as previously described[1]. Sequencing reads were mapped to a custom version of the SS14_v2 reference genome (NC_021508.1) using bwa mem v0.7.17 as previously described[3], generating whole genome multiple sequence alignments using samtools[7] v1.6 and bcftools v1.6, and requiring a minimum of two supporting reads per strand and five in total to call a variant, and a variant frequency/mapping quality cut-off of 0.8. Sites not meeting our filtering criteria were masked to 'N' in the final pseudosequence. We excluded genomes with <75% of genomic positions passing filters at >5x and not masked. Although we have previously shown that this does not substantially affect the tree topology[3], it is notable that samples identified in pairwise analysis as identical over the core genome may have masked sites that would otherwise discriminate them.

#### Phylogenomic analysis

We screened the resulting multiple sequence alignment for recombination using Gubbins[8] v2.4.1, generating whole genome maximum likelihood phylogenies using a K3Pu+F+I model and 10,000 UltraFast bootstraps in IQ-Tree[9] v1.6.10, and inputting SNP-only alignments plus missing constant sites using the `-fconst` flag, as previously described[3]. For analysis of UK genomes in global context, we used a previously described dataset of 526 TPA genomes (including the 237 UK samples studied here, as well as 289 non-UK samples from 21 countries, representing all high quality TPA genomes available at the time of study)[3] with 901 variable sites. For temporal analysis, we used a time-scaled Bayesian maximum credibility tree of 520 global genomes previously generated and evaluated using BEAST2[10] v2.6.3, using a Strict Clock with reference rate prior of $1.23 \times 10^{-7}$ substitutions per site per year, HKY substitution model[11], and Coalescent Bayesian Skyline distribution with 10 populations[11,12]. We extracted UK- and sublineage-specific subtrees from the global analyses using the ape[13] v5.5 and treeio[14] v1.18.1 packages. We used pyjar v0.1.0 (available at https://github.com/simonrharris/pyjar) to perform joint ancestral reconstruction on our maximum likelihood phylogeny, generating a SNP-scaled phylogeny which we used as input into GrapeTree[15] v2.2 to produce minimum spanning trees, where each node represents a cluster of samples separated by a single substitution. All phylogenetic trees were plotted in R[16] v4.1.2 using ggtree[17] v3.2.1.

We performed joint ancestral reconstruction[18] on the maximum likelihood phylogeny using pyjar v0.1.0 (available at https://github.com/simonrharris/pyjar), and extracted the patristic pairwise SNP distance between TPA genomes using the `cophenetic.phylo` function in ape v5.5 to infer pairwise SNP distances between TPA genomes. We constructed single-linkage 'edge list' networks of closely related genomes using the network[19] v1.17.1 package in R, after first filtering all potential sample linkages (edges) to remove those not meeting defined thresholds of pairwise SNP distance (zero SNPs, or less than or equal to two SNPs, dependent on analysis), pairwise temporal distance, and/or sharing the same geographical region. Extraction of network component information was performed using iGraph[20] v1.2.9. Networks were plotted in R using the ggnetwork[21] v0.5.10 and ggplot2[22] v3.3.5 packages. Macrolide resistance alleles were inferred using the competitive mapping approach previously described[1] (available at https://github.com/matbeale/Lihir_Treponema_2020/competitive_mapping_Treponema23S–mod.sh).

We downloaded publicly available map data with Public Health England Region boundaries from the UK Office for National Statistics (https://geoportal.statistics.gov.uk), and used the rgdal[23] v1.5-27, broom[24] v0.7.10 and ggplot2 packages to process and plot map data, and the scatterpie[25] v0.1.7 package to plot pie charts.

#### Supplementary Data

**Supplementary Table 1**. Characteristics of Whole Genome Sequencing (WGS) and all syphilis diagnoses in GUMCAD, England, 2012-2018.

| Variable | WGS (n=237) n | (%) | All diagnoses (n=50,845) n | (%) |
|---|---|---|---|---|
| Year | | | | |
| 2012 | 21 | (8.9) | 4,856 | (9.6) |
| 2013 | 19 | (8.0) | 5,308 | (10.4) |
| 2014 | 12 | (5.1) | 6,342 | (12.5) |
| 2015 | 38 | (16.0) | 7,351 | (14.5) |
| 2016 | 71 | (30.0) | 8,034 | (15.8) |
| 2017 | 48 | (20.3) | 9,177 | (18.0) |
| 2018 | 28 | (11.8) | 9,777 | (19.2) |
| Gender orientation | | | | |
| Heterosexual men (MSW) | 25 | (10.5) | 8,978 | (17.7) |
| GBMSM | 180 | (76.0) | 33,190 | (65.3) |
| Women | 9 | (3.8) | 6,467 | (12.7) |
| Men unknown | 15 | (6.3) | - | - |
| Unknown | 8 | (3.4) | 2,210 | (4.4) |
| Age group | | | | |
| 16-24 years | 30 | (12.7) | 6,194 | (12.2) |
| 25-34 years | 70 | (29.5) | 16,298 | (32.1) |
| 35-44 years | 59 | (24.9) | 13,487 | (26.5) |
| 45+ years | 76 | (32.1) | 14,671 | (28.9) |
| Unknown | 2 | (0.8) | 195 | (0.4) |
| Geography | | | | |
| East Midlands | 5 | (2.1) | 2,398 | (4.7) |
| East of England | 8 | (3.4) | 2,495 | (4.9) |
| London | 118 | (49.8) | 24,326 | (47.8) |
| North East | 24 | (10.1) | 1,497 | (2.9) |
| North West | 6 | (2.5) | 5,079 | (9.5) |
| South East | 29 | (12.2) | 4,776 | (9.4) |
| South West | 15 | (6.3) | 2,082 | (4.1) |
| West Midlands | 7 | (3.0) | 3,763 | (7.4) |
| Yorkshire & Humber | 5 | (2.1) | 2,760 | (5.4) |
| Unknown | 18 | (7.6) | 1,475 | (2.9) |
| UK (Not England) | 2 | (0.8) | 194 | (0.4) |
| UK born | | | | |
| Non-UK born | 66 | (27.8) | 24,191 | (47.6) |
| UK born | 138 | (58.2) | 26,654 | (52.4) |
| Unknown | 33 | (13.9) | - | - |
| HIV status (GBMSM only) | | | | |
| Negative | 121 | (51.1) | 21,888 | (43.1) |
| Positive | 65 | (27.4) | 11,302 | (22.2) |
| Data unavailable | 51 | (21.5) | 17,655 | (34.7) |
| Syphilis stage (matched to diagnoses only, n=140) | | | | |
| Primary | 114 | (81.4) | 14,178 | (27.9) |
| Secondary | 15 | (10.7) | 9,918 | (19.5) |
| Early Latent | 10 | (7.1) | 12,261 | (24.1) |
| Late latent | 1 | (0.7) | 13,032 | (25.6) |
| Cardio | 0 | (0.0) | 635 | (1.3) |
| Neuro | 0 | (0.0) | 821 | (1.6) |

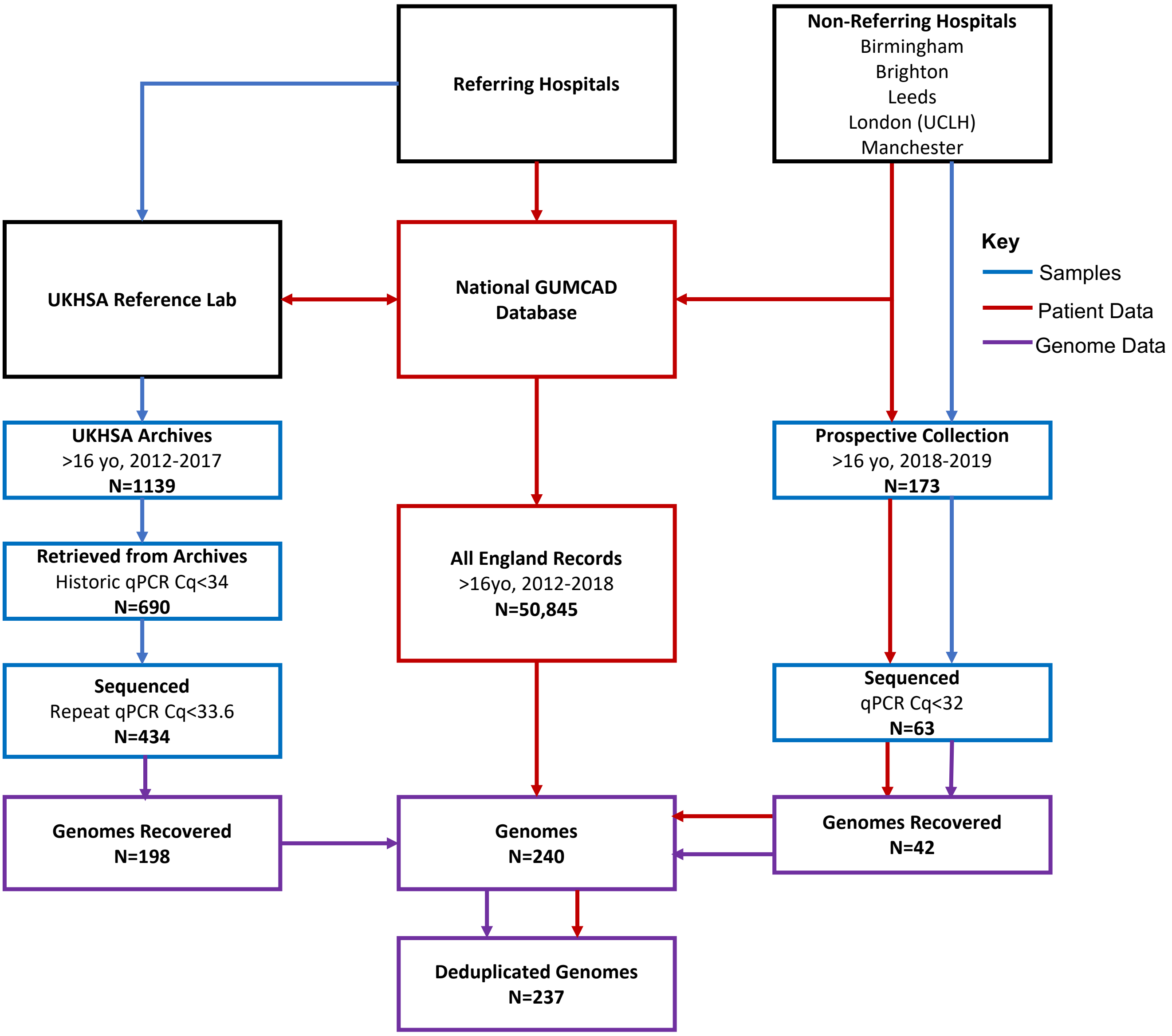


**Supplementary Figure 1. Outline of Samples, Patient data, and Genome data workflows in this study**. Data from all records in the English national STI surveillance system (GUMCAD) from patients >16 years old between 2012-2018 were linked to samples available from the UKHSA archives meeting minimum qPCR thresholds. Separately, we prospectively collected samples from non-referring laboratories (2018-2019) along with metadata from local LIMS, and these were linked back to the records in GUMCAD for deduplication.

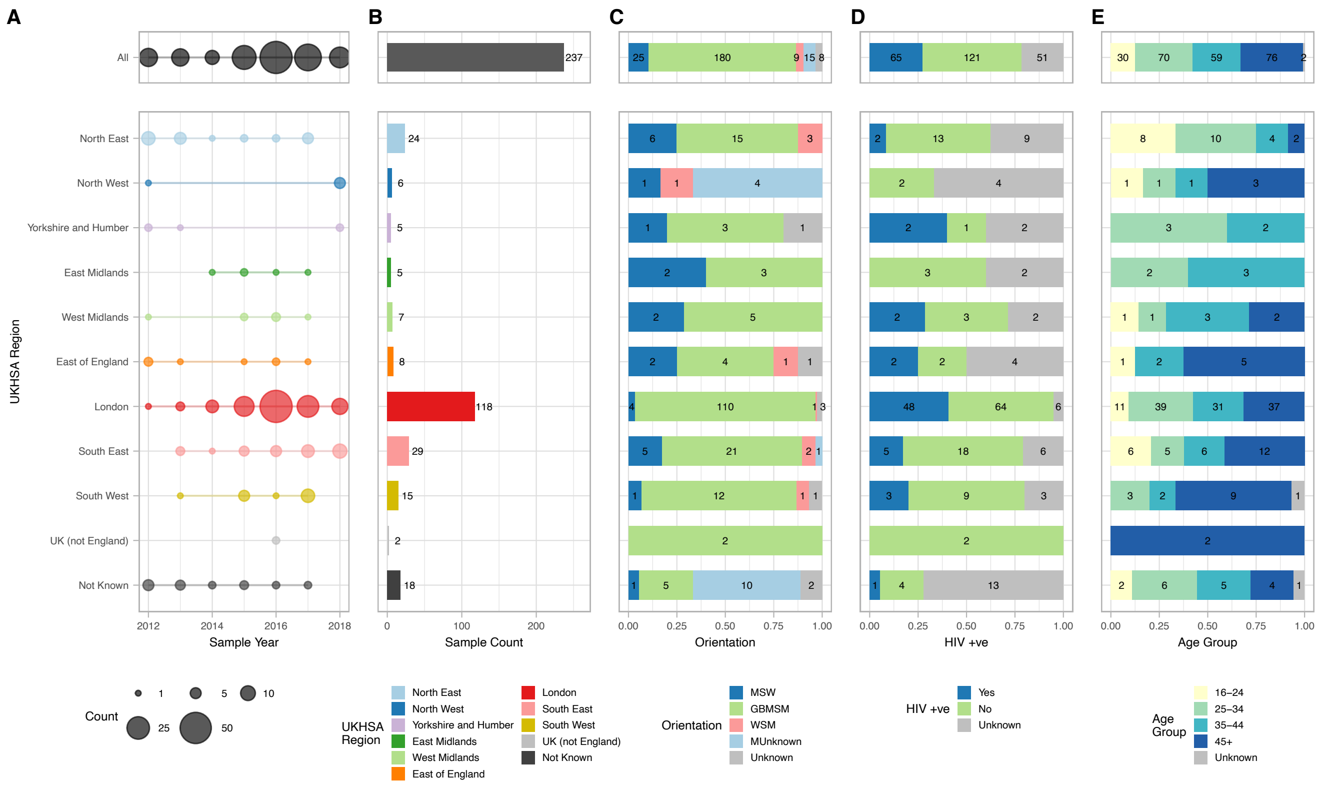

**Supplementary Figure 2. Distribution of all UK genome samples, according to geographical region**. Plot shows collection years (A) and total sample counts (B) per UKHSA Region, then proportion of each group by Gender Orientation (C), HIV Status (D) and Age Group (E). Numbers in bar plots indicate exact sample counts.

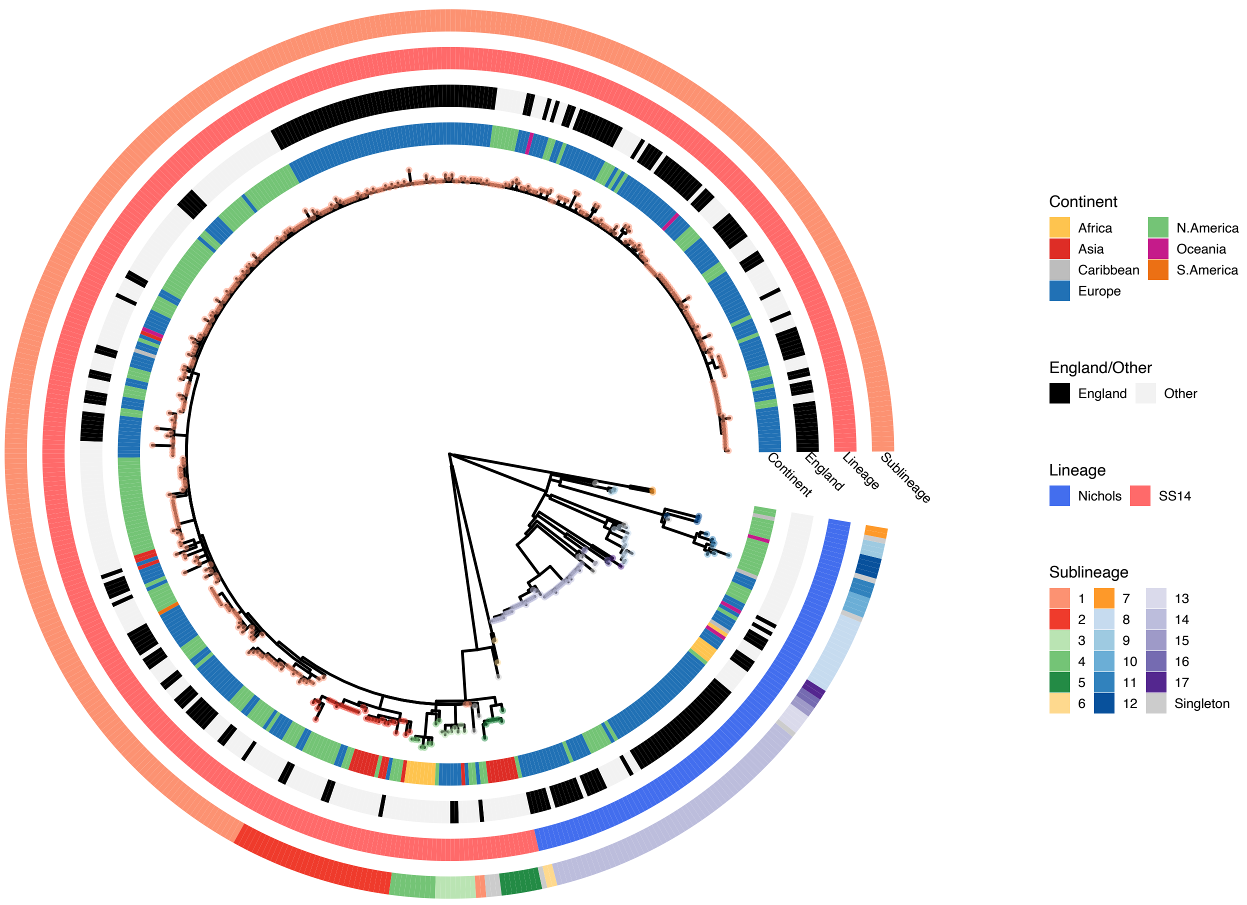


**Supplementary Figure 3. UK genomes are distributed throughout the global phylogeny**. Maximum likelihood phylogeny of 526 genomes from 22 countries, highlighting UK genomes. Tip points are coloured by phylogenetic sublineage (1-17, Singleton), and colour tracks indicate sample continent, whether a sample was collected in the UK, Lineage and Sublineage.

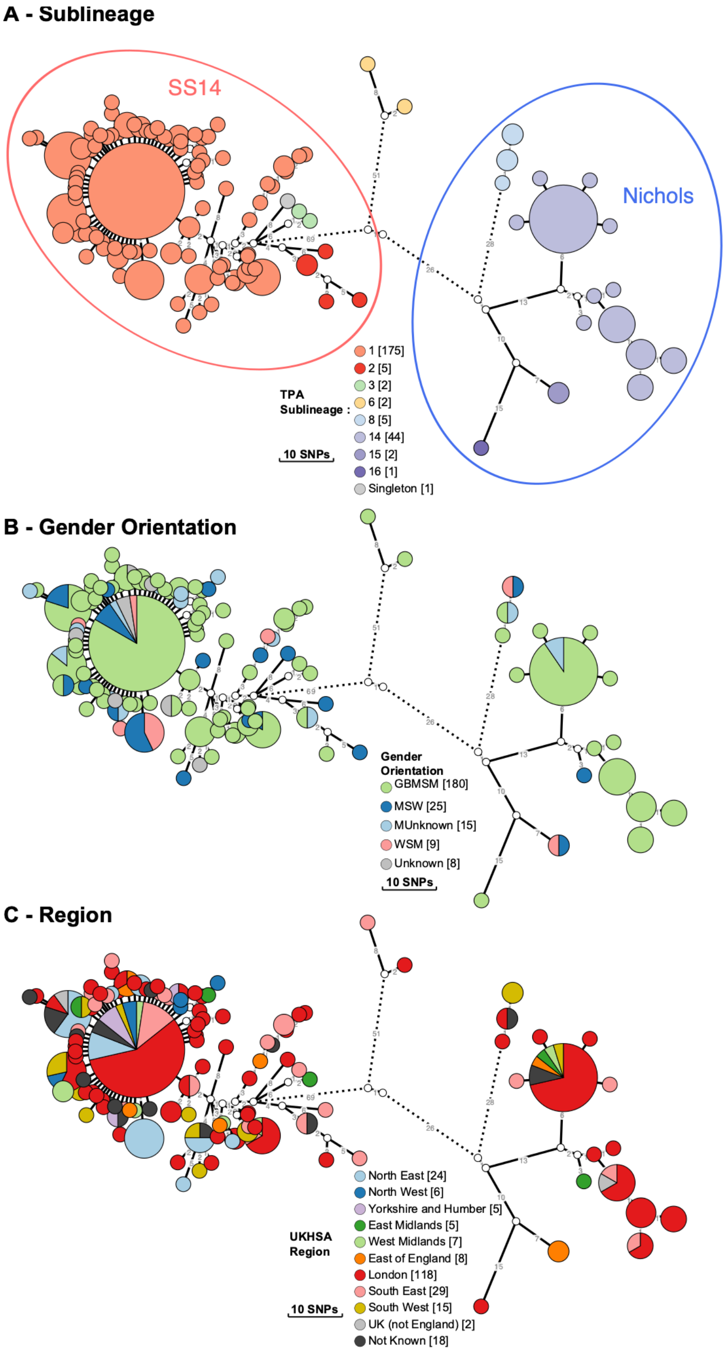


**Supplementary Figure 4. Minimum spanning tree analysis of genomic clustering relationships between samples.** Network visualisation genetic relationships between samples from England. Node size corresponds to the number of identical samples in a cluster, and edge length (with number) to the number of substitutions between clusters (where edges were longer than 12 substitutions, these have been shortened, and this is indicated by dashed lines). Nodes are coloured by proportional membership of each metadata group. A- Clusters coloured by TPA sublineage. B- Clusters coloured by patient gender orientation. C- Clusters coloured by UKHSA Region (England).

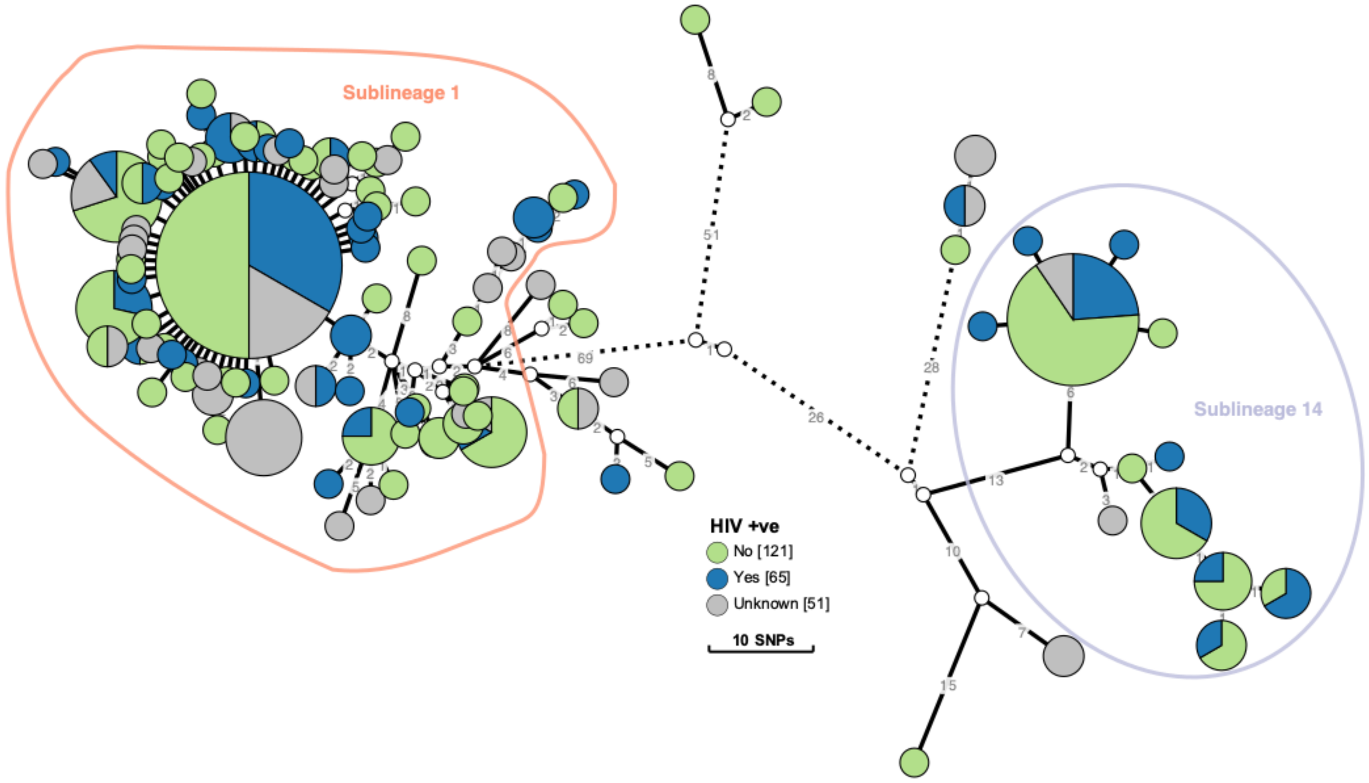


**Supplementary Figure 5. HIV positive individuals are distributed throughout the phylogeny, and are not phylogenetically linked.** Minimum spanning tree network visualisation of genomic clustering relationships between samples from England and HIV status. Node size corresponds to the number of identical samples in a cluster, and edge length (with number) to the number of substitutions between clusters (where edges were longer than 12 substitutions, these have been shortened, and this is indicated by dashed lines). Nodes are coloured by proportion of HIV status, and rings indicate the two common sublineages (1 and 14).

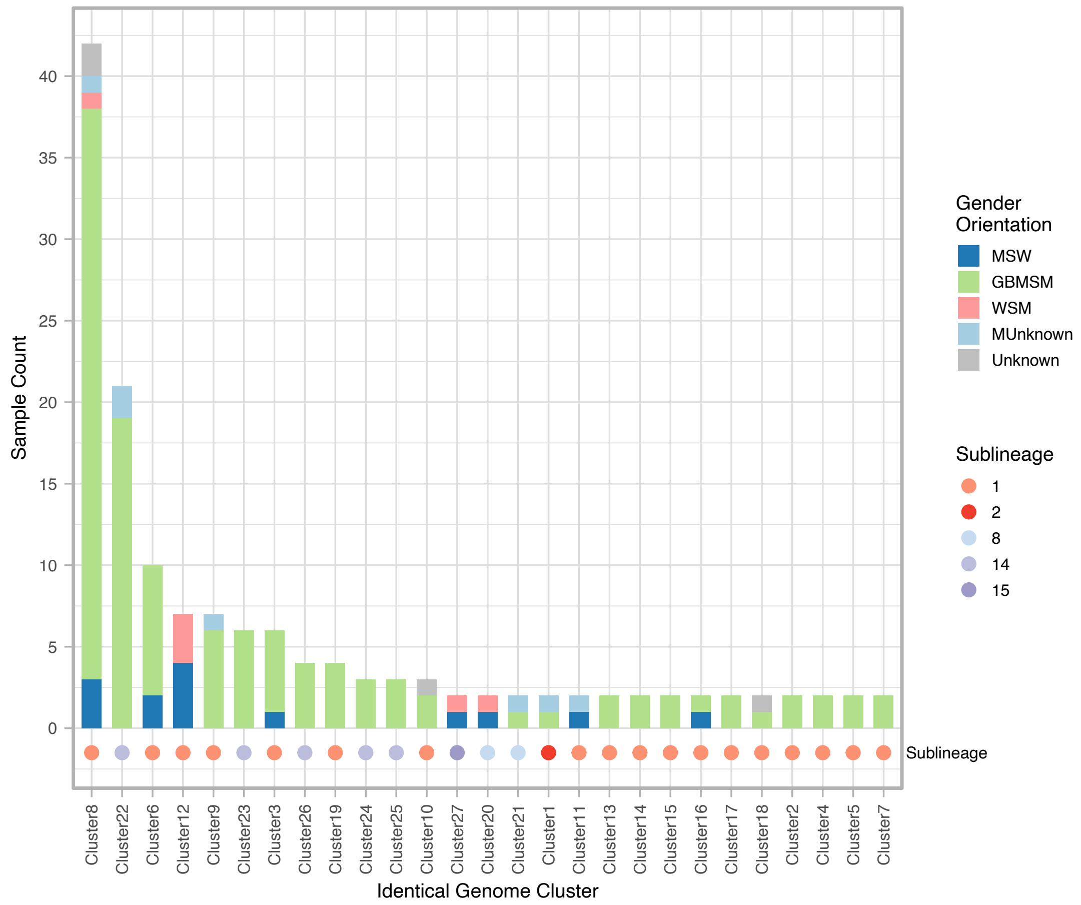


**Supplementary Figure 6. Zero SNP distance clusters show heterosexual transmission networks as well as mixed networks**. Delineation of genomes into clusters separated by zero-pairwise SNPs, indicating sublineage and gender orientation of patients.

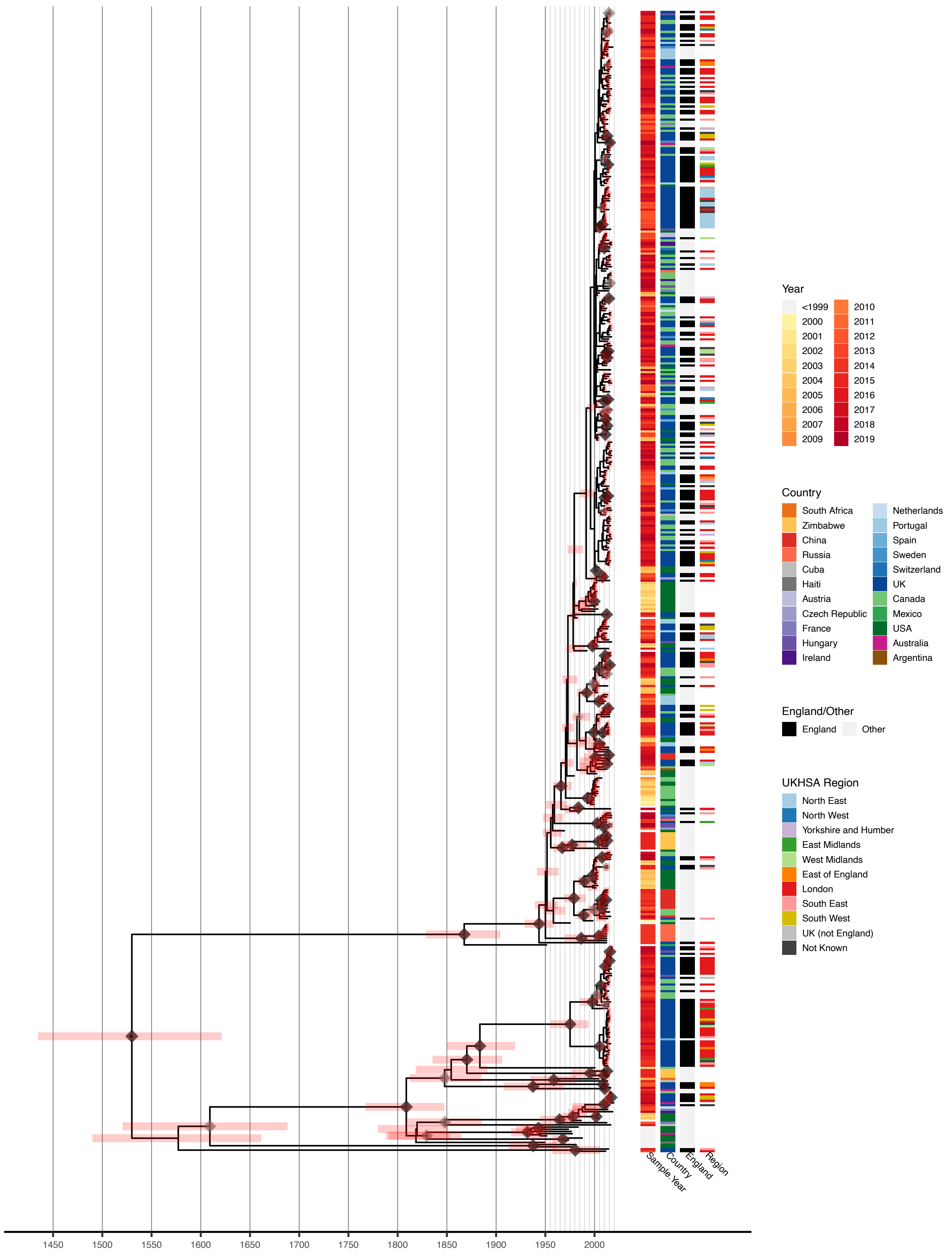


**Supplementary Figure 7. Bayesian time-scaled maximum credibility phylogeny of 526 globally sampled TPA whole genomes**. Colour tracks indicate Sample Collection Year, Country, UK or Other, and UKHSA Region of England.

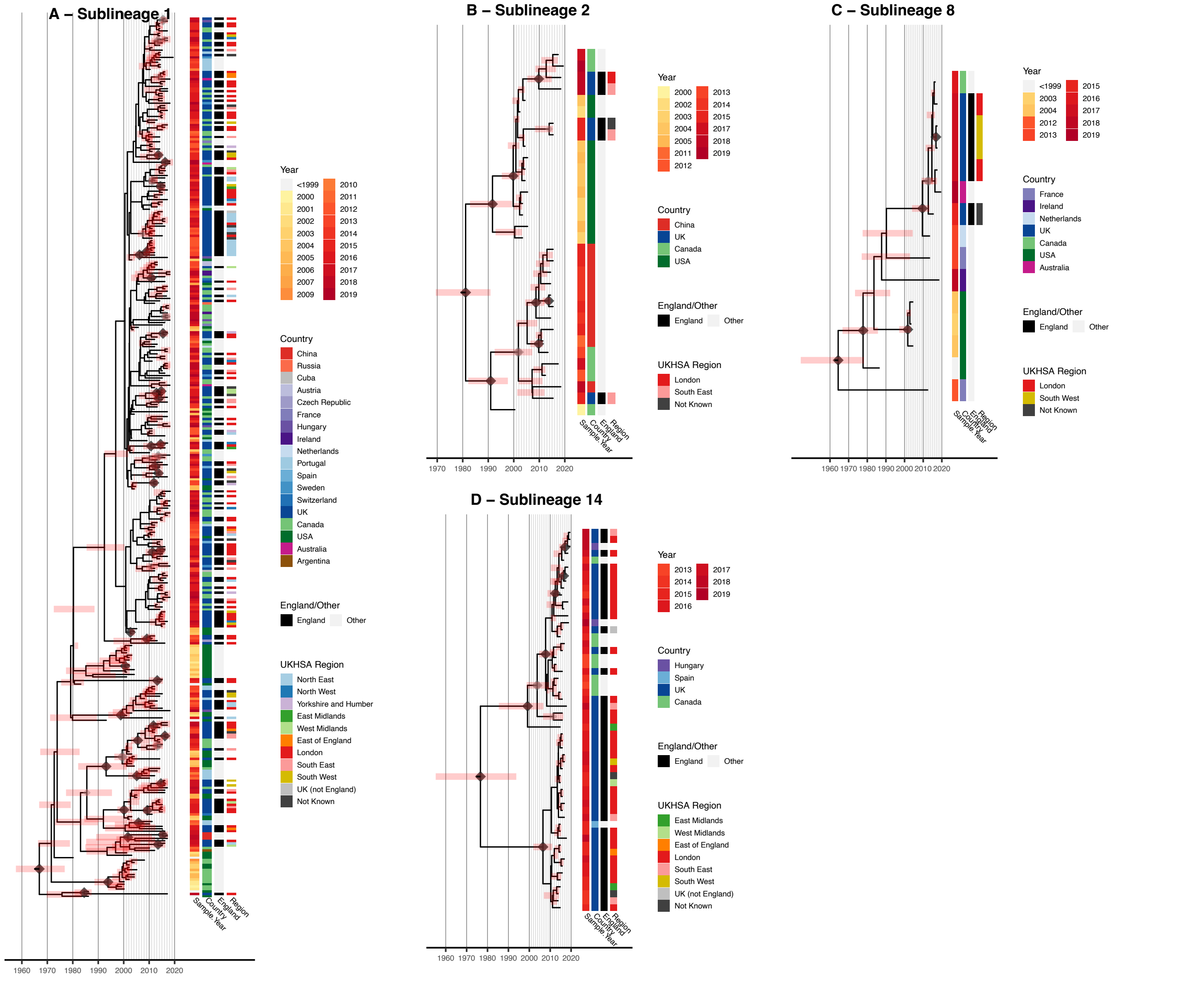


**Supplementary Figure 8. Bayesian phylogenomic and spatiotemporal analysis of major sublineages in England, using UK and Global samples**. Maximum credibility subtree phylogenies derived from a total dataset analysis. A- Sublineage 1, B- Sublineage 2, C- Sublineage 8, D- Sublineage 14. Node points are shaded according to posterior support (black ≥96%, dark grey >91%, light grey >80%). Pink bars on nodes indicate 95% highest posterior density intervals. Colour tracks indicate Sample Collection Year, Country, UK or Other, and UKHSA Region of England.

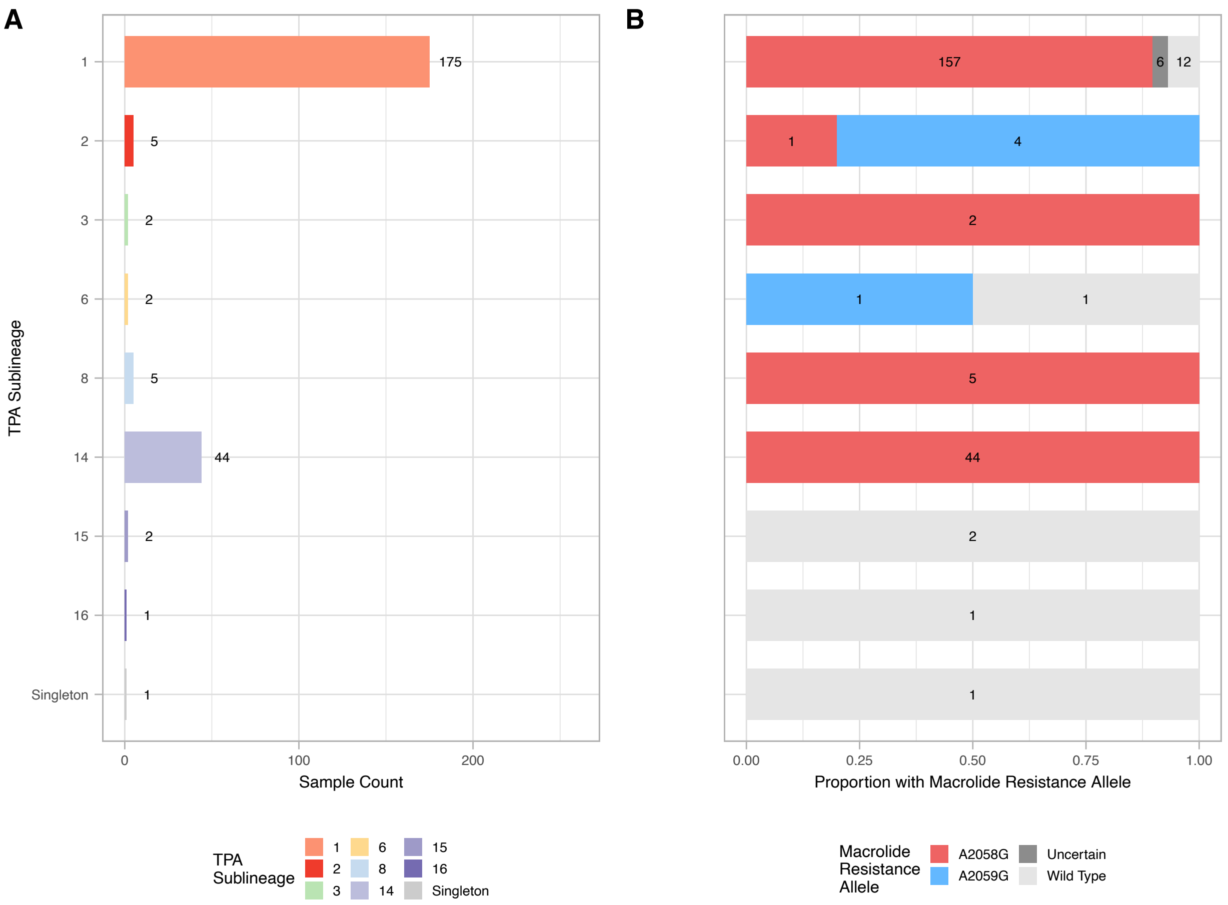


**Supplementary Figure 9. Macrolide resistance alleles are linked to TPA sublineage, with the dominant sublineages circulating in England being resistant**. Proportion of English samples according to 23S macrolide resistance allele (A2058G, A2059G, wild type (sensitive) or uncertain). A – sample count per sublineage, B – proportion of samples with each 23S allele (numbers indicate sample count). Samples with mixed bases or low coverage over the 23S rDNA locus are denoted ‘uncertain’.
